# Self-reported changes in energy balance behaviors during COVID-19 related home confinement: A Cross-Sectional Study

**DOI:** 10.1101/2020.06.10.20127753

**Authors:** Surabhi Bhutani, Jamie A Cooper, Michelle R Vandellen

## Abstract

**Background:** The COVID-19 pandemic has caused people to shelter-at-home for an extended period, resulting in a sudden rise in unstructured time. This unexpected disruption in everyday life has raised concerns about weight management, especially in high-risk populations of women and individuals with overweight and obesity. This study aimed to investigate the changes in behaviors that may impact energy intake and/or energy expenditure in U.S. adults during the home confinement.

**Methods:** Cross-sectional data from 1,779 adults were collected using an online Qualtrics survey between April 24^th^ and May 4^th^, 2020. Self-reported data on demographics, eating behaviors, physical activity, sleep, screen time, takeout food intake, and food purchasing behaviors were collected. Chi-Square analyses were conducted to evaluate differences in the percent of participants reporting increasing, decreasing, or staying the same in each health behavior since the COVID-19 outbreak in their area. Each analysis was followed by comparing whether increases or decreases were more likely for each health behavior. Similar comparisons were made between male and female participants and between body mass index (BMI) categories.

**Results:** We observed an increase in the intake of both healthy and energy-dense unhealthy foods and snacks during the home confinement. Participants also reported increases in sedentary activities and decrease in physical activity, alcohol intake, and consumption of takeout meals during this time. In women, several behavioral changes support greater energy intake and less energy expenditure than men. No clear difference in patterns was observed across BMI status.

**Conclusion:** Acute changes in behaviors underscore the significance of a sudden increase in unstructured time at home on potential weight gain. Our findings support the need to implement and support measures that promote strategies to maintain body weight and establish a methodology to collect body weight data at multiple time points to longitudinally assess the dynamic relationship between behaviors and body weight change.

## INTRODUCTION

The SARS-CoV-2 coronavirus, responsible for the COVID-19 pandemic, has caused significant disruption in the everyday life of people all over the world. In late March 2020, restaurants and shops closed and transitioned to delivery services, and workplaces transitioned to home. In addition, shelter-in-place orders to prevent the spread of the virus were delivered in many cities and states. As a result, people were suddenly confronted with the prospect of sheltering at home for an extended period. One area of great concern with this sudden rise in unstructured time with extended home confinement is its long-term effect on weight management in adults. These concerns are reflected in numerous social media posts referencing “Quarantine 15”, “gaining the COVID-19”, or “fattening the curve”, in the last 3 months. While the fear of excessive weight gain in the general public is palpable, this concern is anecdotal mainly due to the lack of objective scientific evidence. We know from prior literature that small changes in body weight in relatively short periods can become permanent and lead to substantial weight gain over time[1]. Thus, it is imperative to understand the challenges with shelter-in-place practices as they relate to weight management.

Numerous possible challenges can affect energy intake and energy expenditure during this home isolation period, the two components of energy balance[2]. Recent market analyses indicate that people stocked up on energy-dense ultra-processed foods and alcohol, and showed greater interest in cooking/baking high-calorie food[3, 4], possible predecessors to shifting food intake trends. Although these trends provide some evidence of a shift in eating and food purchase behavior, systematic scientific evidence is unavailable to reach any conclusions. Moreover, we do not know whether there is a similar shift for the consumption of healthy foods. Just as shelter-in-place recommendations may shift energy intake behavior, they also likely affect energy expenditure. With people staying indoors (in some cities and states), closures of many local parks and recreation areas, and cancellations of gym and organized sport activities, we suspect a decline in both structured and unstructured activity, and thus reduced energy expenditure. Sedentary activities and screen time are expected to expand; available data already indicates an upward trend in the use of television, internet-connected devices, and applications/web on smart devices[3]. This shift in leisure screen time is possibly displacing physical activity (PA) and contributing to a more sedentary lifestyle[5]. Increased screen time also involves increased exposure to high-frequency blue light and may influence sleep time and sleep quality in adults[6].

To date, no study has systematically examined the health and lifestyle changes as a consequence of COVID-19 related extended home confinement. Therefore, the purpose of this study was to test whether shelter-in-place guidelines—which are associated with alterations to daily routines—may result in changes in behaviors related to both energy intake and energy expenditure. We hypothesized that during shelter-at-home guidelines in place, we would observe an increase in purchase and intake of ultra-processed foods, snacks, and convenience food, an increase in screen time, and a decline in healthy food intake, PA, and sleep time.

## METHODS

### Study Design

This study used a cross-sectional design in which a convenience sample of U.S. adults completed an online survey. The online questionnaire was delivered using the software platform, Qualtrics (Qualtrics® Software Company Provo UT and Seattle WA). An informed consent was obtained from all participants before data collection through the questionnaire. The consent form was presented on the introduction page of the questionnaire, and participants provided their consent by selecting the “I agree to participate” option before they were able to proceed to the questionnaire. The study was approved by the Institutional Review Board at San Diego State University.

### Participants

A total of 1,779 men (43.38%) and women (56.62 %) between the age of 18 and 75 years who had access to the internet, and are living in the U.S, completed the online questionnaire. The Qualtrics questionnaire was administered through either Amazon Mechanical Turk (Mturk) (n=1,267) or via social media, email, and word of mouth (n=511). Mturk (© 2005-2018, Amazon Mechanical Turk, Inc., Seattle, WA) is a web service that enables researchers to survey the target population across the US. Prior studies show that U.S. workers on Mturk are similar to the U.S. population and provide diversity in terms of age, ethnicity, and socioeconomic status, and can provide valid and reliable results for health research[7, 8]. Eligible participants completing the survey through Mturk received a small amount of monetary compensation ($1.66). All participant recruitment and data collection occurred during the 11days from April 24th to May 4^th^, to collect data during peak shelter-in-place guidelines across the country. Participants were primarily Caucasian (77.18%) with additional participants reporting to be Black (6.9%), American Indian or Alaska Native (0.68%), Asian (10.63%), Native Hawaiian (0.25%) or Other (4.35%). A minority of participants identified as Hispanic or Latino (11.35%). Just over half of the participants indicated being employed full-time (55.94%). An additional 15.23% reported being employed part-time. Participants reported being unemployed but in college, full-time (7.58%) or unemployed (9.95%), retired (3.48%), disabled (2.8%), or other (4.23%). Of participants employed full-or part-time, the majority indicated during their work entirely at home (71.50%), while others reported working entire at their usual place of work (17.92%) or a mix of at home and their usual place of work (10.58%).

### Questionnaire

All participants received an electronic invitation to complete the questionnaire through Qualtrics. The total number of questions varied (87 question total) due to follow-up items if a participant answered ‘yes’ vs ‘no’. The questionnaire included the following 7 blocks: demographics, weight behaviors, sleep, and other health behaviors, eating behaviors, PA behaviors, psychological factors, and food purchasing behaviors.

Questions within these categories were aimed at practices and beliefs during the COVID-19 outbreak as well as whether these practices have changed (increased, decreased, or stayed the same). The survey took approximately 25 minutes to complete.

To determine changes in eating behaviors, participants were asked whether their consumption of the following items increased, decreased, or remained the same: fruits, vegetables, caffeine, drinking non-diet drinks that included all sugar sweetened beverages (SSB) (for example, Coke, Pepsi, flavored juice drinks, sports drinks, sweetened teas, coffee drinks, energy drinks, electrolyte replacement drinks), and drinking diet soda or other diet drinks. To determine a change in consumption of processed and ultra-processed foods, a list of foods were presented as described in the NOVA classification system which groups foods according to the extent and purpose of industrial processing[9]. Additionally, we collected information on the change in consumption of following foods generally consumed as snacks (cake, cookie, ice-cream, other desserts; chips, popcorn, pretzels, and crackers; gummy snacks, fruit candy, sour gummy, or other fruity candies; fruit; vegetables; chocolate; yogurt/cheese). Change in consumption of restaurant/take-out/fast food/delivery food and alcohol intake was also recorded.

To determine PA, the International Physical Activity Questionnaire (IPAQ short version)[10], which is valid and reliable[11, 12], was incorporated. The IPAQ uses metabolic equivalents (METs) to represent the energy exerted through PA. Detailed scoring for the different PA categories has been described previously[13]. Additional questions about whether sitting, walking, moderate, or vigorous PA had changed during the COVID-19 outbreak in their area were also assessed using “I am doing more”, “I am doing the same”, and “I am doing less” options. Change in screen time was determined by asking questions on change in time spent on watching television, social media, or other leisurely activities such as video games, computer, email etc. since COVID-19 outbreak. Sleep quality was determined by the average number of hours spent on sleeping each night and whether the sleep hours are more, the same, or less since the outbreak, compared to before the outbreak. To assess sleep quality, we used the validated Stanford Sleepiness Scale to rate the alertness of the participant after waking up in the morning[14].

Finally, we collected information on food purchase behavior. In particular, we asked how much participants endorsed 1) worrying that they will not be able to purchase food or will run out of food, 2) making purchase decisions based on how longs things will last, 3) making food purchase decisions based on what is available in the grocery store, 4) making food purchase decisions based on what foods are healthier, 5) purchasing more alcoholic beverages. The ratings were made on a 7-point Likert scale (1= strongly disagree; 7=strongly agree). Finally, participants were asked to rank-order five considerations for purchasing food and drink (i.e., nutrition content, availability, expiration date/shelf life, desire/craving, and cost).

### Data analysis

SAS version 9.4 (Cary, NC) was used for all statistical analysis, and significance was set two-tailed at p<0.05. Chi Square analyses were conducted to evaluate differences in percent of participants reporting increasing, decreasing, or staying the same in each health behavior. We followed each analysis with a comparison among people who reported changing, comparing whether increases or decreases were more likely. After examining these main effects, we compared patterns of change across sex (male vs. female) and across body mass index (BMI) group (underweight, normal weight, overweight, obese). We then compared whether, among each subgroup (e.g., males, females, normal weight, obese), individuals who changed were more likely to report increasing or decreasing a given health behavior. Additional analyses examined reasons for food purchase behavior.

## RESULTS

A total of 1,779 participants initially completed the questionnaire. One hundred and seventy participants were removed from data analysis due to poor data quality, for a final study sample of n=1,609 (Figure 1). Subject characteristics and comparison of health-related measures between men and women can be found in Table 1. Approximately 57% of the study sample were women and 43% were men. There were no differences between men and women for average age, hours of sleep, or, BMI. Men did report high Kcal METs and moderate METs. Conversely, women reported poor sleep quality and had higher total weekly METs and Vigorous METs compared to men.

**Table 1.**
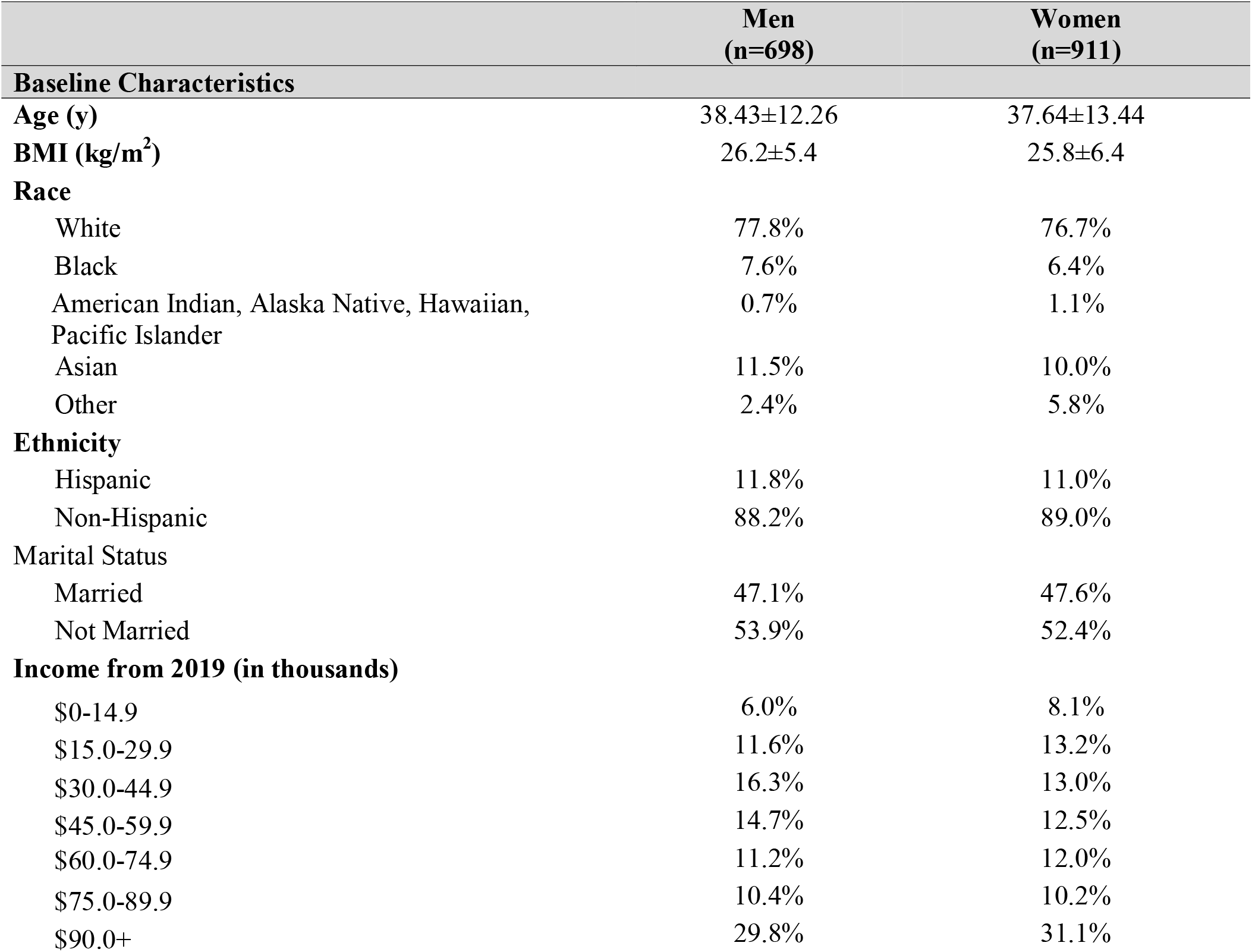
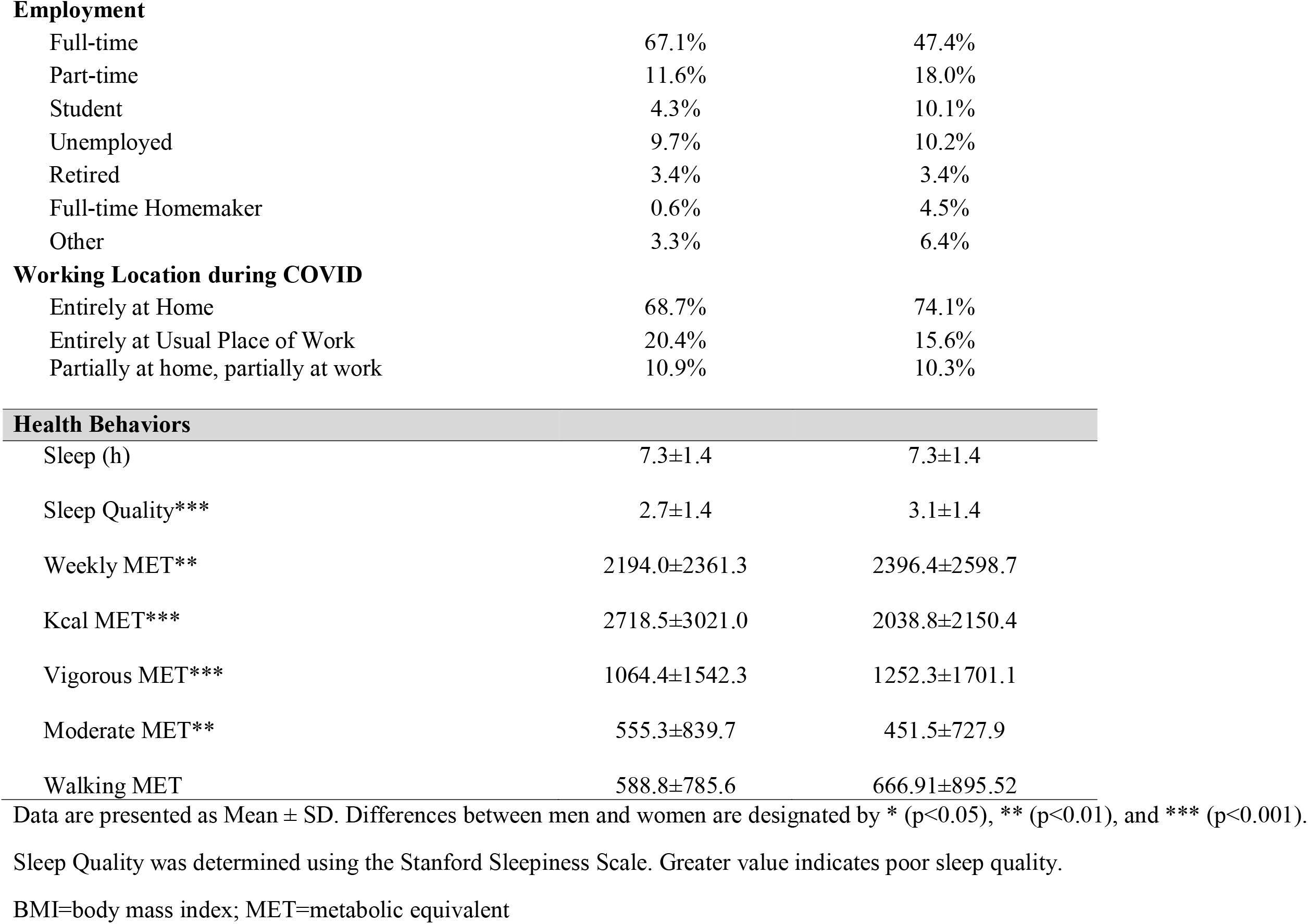
Baseline Characteristics and Health Behavior Measures by Sex

| **Baseline Characteristics** | **Men**<br>**(n=698)** | **Women**<br>**(n=911)** |
| --- | --- | --- |
| **Age (y)** | 38.43±12.26 | 37.64±13.44 |
| **BMI (kg/m<sup>2</sup>)** | 26.2±5.4 | 25.8±6.4 |
| **Race** |  |  |
| White | 77.8% | 76.7% |
| Black | 7.6% | 6.4% |
| American Indian, Alaska Native, Hawaiian,<br>Pacific Islander | 0.7% | 1.1% |
| Asian | 11.5% | 10.0% |
| Other | 2.4% | 5.8% |
| **Ethnicity** |  |  |
| Hispanic | 11.8% | 11.0% |
| Non-Hispanic | 88.2% | 89.0% |
| **Marital Status** |  |  |
| Married | 47.1% | 47.6% |
| Not Married | 53.9% | 52.4% |
| **Income from 2019 (in thousands)** |  |  |
| \$0-14.9 | 6.0% | 8.1% |
| \$15.0-29.9 | 11.6% | 13.2% |
| \$30.0-44.9 | 16.3% | 13.0% |
| \$45.0-59.9 | 14.7% | 12.5% |
| \$60.0-74.9 | 11.2% | 12.0% |
| \$75.0-89.9 | 10.4% | 10.2% |
| \$90.0+ | 29.8% | 31.1% |
| **Employment** |  |  |
| Full-time | 67.1% | 47.4% |
| Part-time | 11.6% | 18.0% |
| Student | 4.3% | 10.1% |
| Unemployed | 9.7% | 10.2% |
| Retired | 3.4% | 3.4% |
| Full-time Homemaker | 0.6% | 4.5% |
| Other | 3.3% | 6.4% |
| **Working Location during COVID** |  |  |
| Entirely at Home | 68.7% | 74.1% |
| Entirely at Usual Place of Work | 20.4% | 15.6% |
| Partially at home, partially at work | 10.9% | 10.3% |
| **Health Behaviors** |  |  |
| Sleep (h) | 7.3±1.4 | 7.3±1.4 |
| Sleep Quality*** | 2.7±1.4 | 3.1±1.4 |
| Weekly MET** | 2194.0±2361.3 | 2396.4±2598.7 |
| Kcal MET*** | 2718.5±3021.0 | 2038.8±2150.4 |
| Vigorous MET*** | 1064.4±1542.3 | 1252.3±1701.1 |
| Moderate MET** | 555.3±839.7 | 451.5±727.9 |
| Walking MET | 588.8±785.6 | 666.91±895.52 |

Data are presented as Mean ± SD. Differences between men and women are designated by \* (p<0.05), \*\* (p<0.01), and \*\*\* (p<0.001).
Sleep Quality was determined using the Stanford Sleepiness Scale. Greater value indicates poor sleep quality.
BMI=body mass index; MET=metabolic equivalent

**Figure 1.**
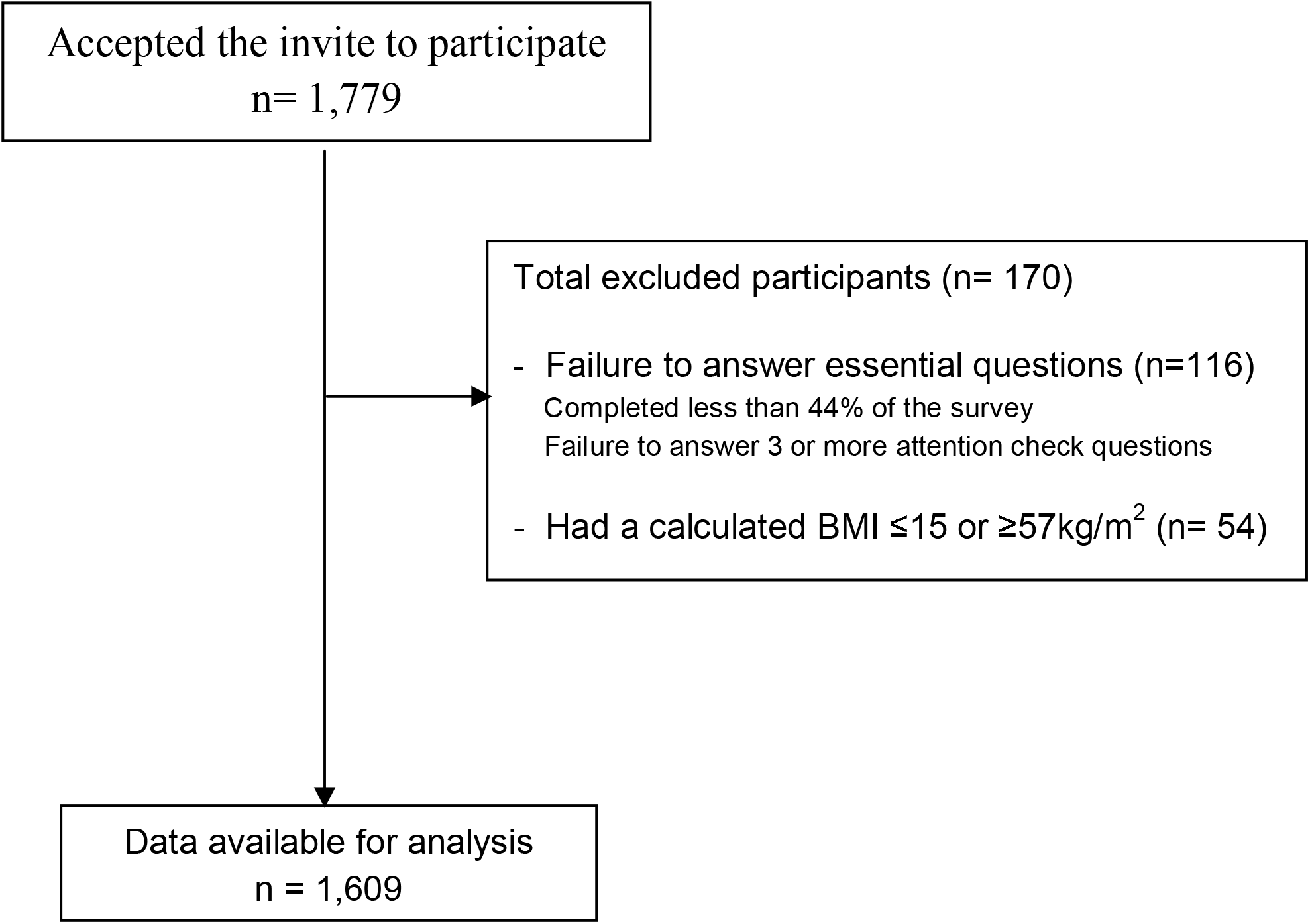
Flow chart of study participants based on the STrengthening the Reporting of OBservational studies in Epidemiology (STROBE) guidelines

### Changes in Eating Behavior

We observed significant differences in the proportion of participants reporting increasing, decreasing, or staying the same for different types or categories of food consumption (Table 2). Consumption of vegetables, fruit, SSBs, processed foods, and ultra-processed foods was more likely to increase than decrease. With regards to snacks, intake of chocolate, fruits, vegetables, chips (or similar salty snacks), dessert, and yogurt/cheese was more likely to increase than decrease. Consumption of gummy/fruity candies and diet soda was equally likely to increase or decrease. No foods were more likely to be reported as having decreased consumption relative to increased consumption.

**Table 2.** Self-Reported Changes in Food Consumption Since the COVID-19 Outbreak

| Consumption Change | Increase | Same | Decrease | Chi Square of Different Patterns | Chi Square of Increase vs. Decrease |
| --- | --- | --- | --- | --- | --- |
| Vegetables | 18.96 | 65.02 | 16.02 | 707.43*** | 3.88* |
| Fruits | 21.25 | 60.95 | 17.8 | 530.88*** | 4.67* |
| Non-diet drinks (all SSBs) | 16.39 | 71.41 | 12.20 | 875.03*** | 8.21** |
| Diet Soda or other diet drinks | 12.13 | 74.35 | 13.51 | 880.24*** | 0.86 |
| Processed foods | 31.76 | 55.79 | 12.45 | 424.91*** | 126.66*** |
| Ultra-processed foods | 24.62 | 59.81 | 15.57 | 477.79*** | 29.73*** |
| Foods as Snacks |  |  |  |  |  |
| Chocolate | 24.00 | 61.75 | 14.25 | 519.07*** | 34.14*** |
| Fruit (Snack) | 26.04 | 57.34 | 16.62 | 504.01*** | 30.91*** |
| Vegetable (Snack) | 20.29 | 64.02 | 15.70 | 622.73*** | 8.55** |
| Gummy/Fruity candies | 16.56 | 66.42 | 17.02 | 528.65*** | 0.07 |
| Chips/popcorn/pretzels/crackers | 36.38 | 51.58 | 12.04 | 347.17*** | 178.01*** |
| Desserts (Cake/cookie/ice-cream) | 31.39 | 55.38 | 13.23 | 381.17*** | 104.99*** |
| Yogurt/Cheese | 19.08 | 71.17 | 9.76 | 895.67*** | 41.04*** |

\* Denotes statistical significance at \*(p < 0.05), \*\* (p < 0.01) or, \*\*\* (p < 0.001).
SSBs = sugar sweetened beverages

### Changes in Health Behaviors

As Table 3 shows, we observed differences in the proportion of people who indicated engaging in each behavior more, less, or the same since the COVID-19 outbreak. Participants were more likely to report increases (vs. decreases) in hours of sleep, smoking, consumption of caffeine, leisure screen (i.e., computer/phone) time, television viewing time, and sitting time. Conversely, participants were more likely to report decreases (vs. increases) in vigorous PA, moderate PA, walking, drinking alcohol, and consuming take-out/fast food. The largest effects were observed for takeout/restaurant food such that participants were more likely to decrease restaurant food consumption than increase it, and for sedentary behaviors such that participants were more likely to increase than to decrease sedentary behaviors (i.e., sitting, TV time, screen time).

**Table 3.** Self-Reported Changes in Health Behaviors Since the COVID-19 Outbreak

|  | **Increase** | **Same** | **Decrease** | **Chi Square of**<br>**Different Patters** | **Chi Square of**<br>**Increase vs. Decrease** |
| --- | --- | --- | --- | --- | --- |
| Sleep | 27.47 | 52.69 | 19.84 | 283.43*** | 19.69*** |
| Smoking | 31.95 | 54.77 | 13.28 | 62.45*** | 18.58*** |
| Alcohol Intake | 19.05 | 56.25 | 24.7 | 376.02*** | 11.35*** |
| Takeout/Fast food | 11.61 | 29.00 | 59.39 | 553.00*** | 506.71*** |
| Caffeine intake | 20.21 | 67.97 | 11.81 | 871.48*** | 34.89*** |
| Leisure Screen Time | 53.92 | 41.21 | 4.88 | 605.43*** | 637.22*** |
| TV Time | 50.55 | 43.99 | 5.47 | 553.44*** | 564.18*** |
| Sitting Time | 58.45 | 35.90 | 5.65 | 655.57*** | 677.04*** |
| Walking | 29.15 | 35.69 | 35.17 | 12.25** | 8.71** |
| Moderate PA | 17.56 | 50.16 | 32.27 | 246.74*** | 67.01*** |
| Vigorous PA | 19.02 | 45.78 | 35.20 | 169.00*** | 74.91*** |

\* Denotes statistical significance at \*(p < 0.05), \*\* (p < 0.01) or, \*\*\* (p < 0.001).
PA=physical activity; TV=television.

### Sex Differences in Eating Behavior Changes during the COVID-19 Outbreak

We also observed sex differences in changes in eating behaviors during the pandemic (Table 4). We showed that men and women differed in patterns of change for consumption of all foods but soft drinks. For consumption of fruit, processed foods, and ultra-processed foods, females were more likely to report increases (vs. decreases) than males. Males were more likely to report decreases than increases in diet soft drinks whereas females were more likely to report increases than decreases in regular SSBs. Females were also more likely than males to report increases (vs. decreases) in snacking on chocolate, fruits, vegetables, chips, desserts, and yogurt/cheese.

**Table 4.** Comparison of Patterns of Change in Food Consumption Across Sex Since the COVID-19 Outbreak

| Consumption Change |  | Increase | Same | Decrease | Chi Square of Different Patterns Across Sex | Chi Square of Increase vs. Decrease |
| --- | --- | --- | --- | --- | --- | --- |
| Vegetables | Males | 15.51 | 70.46 | 14.03 | 15.88** | 0.50 |
|  | Females | 21.61 | 60.86 | 17.53 |  | 3.75 |
| Fruits | Males | 17.32 | 65.66 | 17.02 | 13.14** | 0.02 |
|  | Females | 24.23 | 57.37 | 18.40 |  | 6.97*** |
| Non-diet drinks (all SSBs) | Males | 13.68 | 74.19 | 12.14 | 5.82 | 0.54 |
|  | Females | 18.51 | 69.24 | 12.25 |  | 9.56** |
| Diet Soda or other diet drinks | Males | 7.85 | 76.64 | 15.51 | 18.63*** | 13.45*** |
|  | Females | 15.79 | 72.41 | 11.80 |  | 3.61 |
| Processed foods | Males | 23.56 | 63.83 | 12.61 | 38.39*** | 21.78*** |
|  | Females | 38.15 | 49.53 | 12.32 |  | 111.56*** |
| Ultra-processed foods | Males | 19.97 | 63.81 | 16.22 | 13.44** | 2.48 |
|  | Females | 28.27 | 56.67 | 15.06 |  | 32.95*** |
| Foods as Snacks |  |  |  |  |  |  |
| Chocolate | Males | 17.22 | 68.23 | 14.55 | 27.51*** | 1.35 |
|  | Females | 29.21 | 56.76 | 14.03 |  | 41.44*** |
| Fruit (Snack) | Males | 22.38 | 63.58 | 14.04 | 18.34*** | 12.36*** |
|  | Females | 28.88 | 52.51 | 18.62 |  | 18.58*** |
| Vegetable (Snack) | Males | 17.71 | 68.03 | 14.26 | 8.04* | 2.37 |
|  | Females | 22.29 | 60.90 | 16.81 |  | 6.31* |
| Gummy/Fruity candies | Males | 13.19 | 69.69 | 17.13 | 8.04* | 2.60 |
|  | Females | 19.49 | 63.59 | 16.92 |  | 1.06 |
| Chips/popcorn/pretzels/crackers | Males | 30.91 | 56.62 | 12.46 | 14.95*** | 49.78*** |
|  | Females | 40.61 | 47.68 | 11.71 |  | 130.93*** |
| Desserts (Cake/cookie/ice-cream) | Males | 22.56 | 62.18 | 15.26 | 39.40*** | 8.69** |
|  | Females | 38.14 | 50.19 | 11.68 |  | 113.14*** |
| Yogurt/Cheese | Males | 14.62 | 75.42 | 9.97 | 14.01*** | 5.30* |
|  | Females | 22.60 | 67.81 | 9.59 |  | 40.00*** |

\* Denotes statistical significance at \*(p < 0.05), \*\* (p < 0.01) or, \*\*\* (p < 0.001).
SSBs = sugar sweetened beverages

### Sex Differences in Health Behavior Changes during the COVID-19 Outbreak

Similar to changes in eating behaviors between sexes, some changes in health behavior during COVID-19 also differed by sex. As Table 5 shows, patterns of change in sleep time, takeout food, TV time, screen time, sitting time, walking PA, and moderate PA differed for males and females. Changes in smoking, alcoholic drink consumption, caffeine, and vigorous PA activity were similar for men and women. The likelihood of increasing (vs. decreasing) sleep and sedentary behaviors (i.e., sitting time, TV time, screen time) was larger for women than men. In contrast, men were more likely to report decreases (vs. increases) in walking, moderate PA, and vigorous PA than women.

**Table 5.**
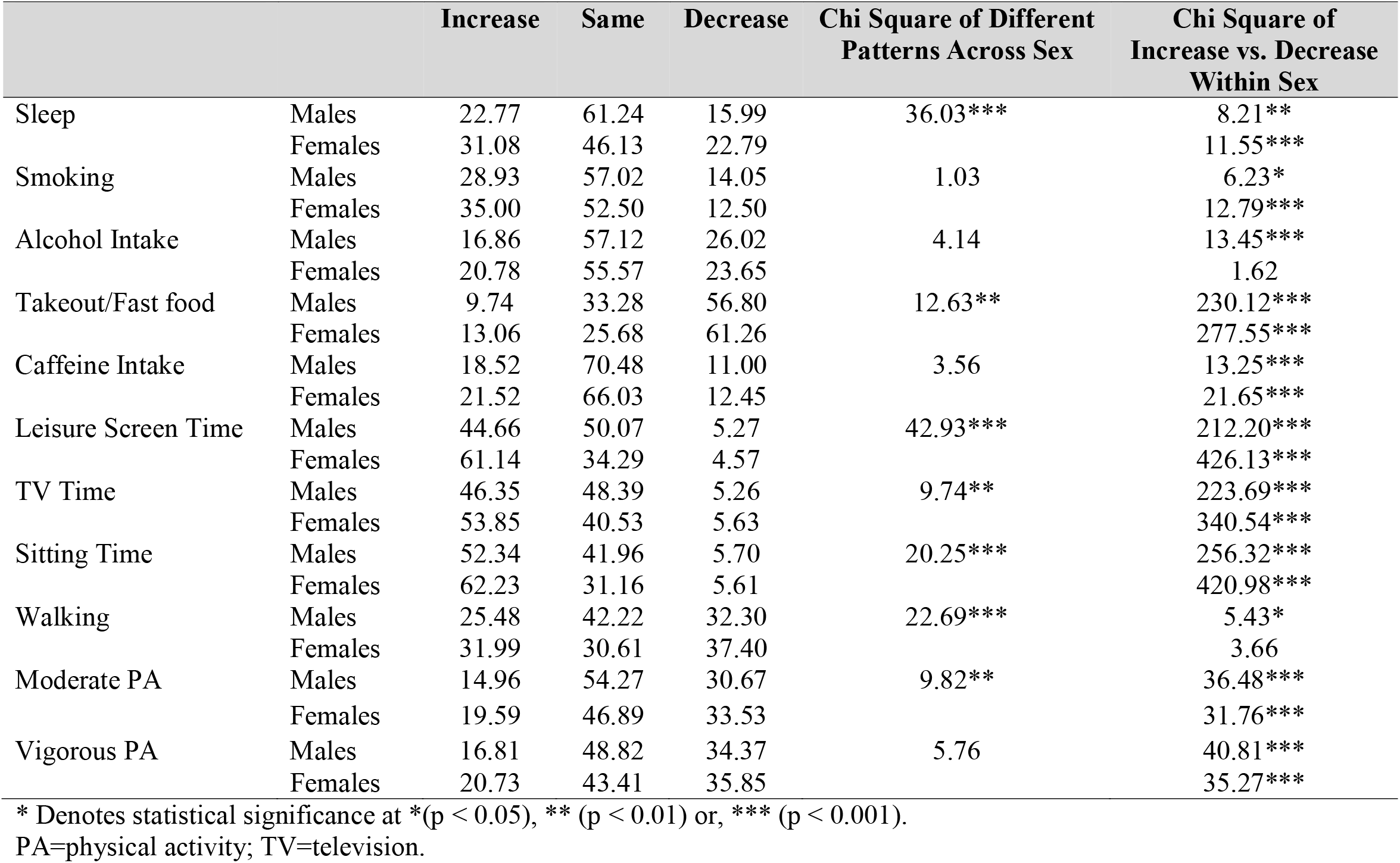
Comparison of Patterns of Change in Health Behaviors Across Sex Since the COVID-19 Outbreak

|  |  | Increase | Same | Decrease | Chi Square of Different<br>Patterns Across Sex | Chi Square of<br>Increase vs. Decrease<br>Within Sex |
| --- | --- | --- | --- | --- | --- | --- |
| Sleep | Males | 22.77 | 61.24 | 15.99 | 36.03*** | 8.21** |
|  | Females | 31.08 | 46.13 | 22.79 |  | 11.55*** |
| Smoking | Males | 28.93 | 57.02 | 14.05 | 1.03 | 6.23* |
|  | Females | 35.00 | 52.50 | 12.50 |  | 12.79*** |
| Alcohol Intake | Males | 16.86 | 57.12 | 26.02 | 4.14 | 13.45*** |
|  | Females | 20.78 | 55.57 | 23.65 |  | 1.62 |
| Takeout/Fast food | Males | 9.74 | 33.28 | 56.80 | 12.63** | 230.12*** |
|  | Females | 13.06 | 25.68 | 61.26 |  | 277.55*** |
| Caffeine Intake | Males | 18.52 | 70.48 | 11.00 | 3.56 | 13.25*** |
|  | Females | 21.52 | 66.03 | 12.45 |  | 21.65*** |
| Leisure Screen Time | Males | 44.66 | 50.07 | 5.27 | 42.93*** | 212.20*** |
|  | Females | 61.14 | 34.29 | 4.57 |  | 426.13*** |
| TV Time | Males | 46.35 | 48.39 | 5.26 | 9.74** | 223.69*** |
|  | Females | 53.85 | 40.53 | 5.63 |  | 340.54*** |
| Sitting Time | Males | 52.34 | 41.96 | 5.70 | 20.25*** | 256.32*** |
|  | Females | 62.23 | 31.16 | 5.61 |  | 420.98*** |
| Walking | Males | 25.48 | 42.22 | 32.30 | 22.69*** | 5.43* |
|  | Females | 31.99 | 30.61 | 37.40 |  | 3.66 |
| Moderate PA | Males | 14.96 | 54.27 | 30.67 | 9.82** | 36.48*** |
|  | Females | 19.59 | 46.89 | 33.53 |  | 31.76*** |
| Vigorous PA | Males | 16.81 | 48.82 | 34.37 | 5.76 | 40.81*** |
|  | Females | 20.73 | 43.41 | 35.85 |  | 35.27*** |

\* Denotes statistical significance at \*(p < 0.05), \*\* (p < 0.01) or, \*\*\* (p < 0.001).
PA=physical activity; TV=television.

### Food Purchase Behavior

As Table 6 shows, participants overall reported low agreement that they were worrying about running out of being able to purchase food, low agreement that they were making food purchases based on health, and low agreement that they were purchasing more alcohol. They indicated high agreement that they were purchasing food based on how long it would last and what was available. We compared the three items assessing reasons for purchase behavior using a repeated measures ANOVA. This analysis indicated a main effect of reason, *F* (2, 2864) = 374.95, p <.001, suggesting that availability was the reason people most agreed with as a reason for their purchase behavior and healthiness was the least likely reason to influence purchase behavior. People reported their top consideration for food purchase behavior to be nutrition (23.44%), availability (20.35%), expiration date/shelf life (20.01%), desire/craving (19.95%), and then cost (16.25%), X^2^ (4) = 19.35, p < .001. The only sex difference in food purchase behavior was that females were more likely to indicate purchasing food based on what was available than males.

**Table 6.** Comparison of Food Purchase Behaviors in the Total Sample and by Sex Since the COVID-19 Outbreak

|  | **Total Sample**<br>**(n = 1439)** | **Men**<br>**(n=660)** | **Women**<br>**(n=779)** |
| --- | --- | --- | --- |
| Worry about not being able to purchase/running out of food | 3.29±1.89*** | 3.27±1.86 | 3.30±1.92 |
| Purchasing food based on how long it will last | 4.29±1.82*** | 4.20±1.78 | 4.36±1.85 |
| Purchasing based on what is available | 4.89±1.68*** | 4.6±1.69 | 5.08±1.64 |
| Purchasing based on what foods are healthier | 3.43±1.70*** | 3.39±1.70 | 3.47±1.70 |
| Purchasing more alcoholic beverages | 2.61±1.99*** | 2.68.0±1.99 | 2.55±1.99 |

Data are presented as Mean ± SD. For total sample, significance refers to comparisons between the sample agreement and the scale midpoint (4). Differences between men and women are designated by \* (p<0.05), \*\* (p<0.01), and \*\*\* (p<0.001) following the question content. Note that total sample size differs because not all participants completed the survey.

### Weight Status related Changes in Behaviors Since the COVID-19 Outbreak

We observed differences in eating behaviors and health behaviors by weight status. Data shown in Supplementary Table. 1 suggests that BMI was significantly and negatively associated with sleep and all measures of PA except for moderate PA. Sleep quality did not differ between BMI groups. With regards to snacking pattern, underweight participants were less likely to report increases than decreases in snacking, and underweight and normal weight participants were less likely to report increases than decreases in pre-packaged food. No other differences in eating behaviors were reported between BMI categories (Supplementary Table. 2). Similar to eating behaviors, there were far fewer differences in health behavior changes during the pandemic by weight status (Supplementary Table. 3). The only group to demonstrate differences in likelihood of reporting increases vs. decreases in sleep was the normal weight group (increase > decrease). We also observed different patterns of change for caffeine consumption such that all groups but underweight were more likely to report increases than decreases. Underweight people were also unlikely to report the decrease (vs. increase) in moderate PA observed in all other weight groups. With regards to food purchasing behaviors, underweight individuals were more likely to report worry about being able to purchase food to purchase foods based on what was healthier, and to purchase more alcoholic beverages (Supplementary Table. 4).

## DISCUSSION

Small changes in body weight in relatively short periods may lead to substantial weight gain over time[1, 15]. Notably, prior research suggests that short bouts of weight fluctuation, such as during the winter holiday period (November to January), may contribute to half of annual weight gain[16-18]. This rapid weight gain is likely to remain, especially in the high-risk populations of women and individuals with overweight and obesity[17, 19, 20]. Considering the possibility that extended home confinement due to COVID-19 may have a similar effect on body weight, in this study, we aimed to report alterations in behaviors that may impact energy intake and energy expenditure in U.S. adults. We observed an increase in the intake of both healthy and energy-dense unhealthy foods and snacks during the home confinement. Participants also reported increases in sedentary activities and decreases in PA, alcohol intake, and consumption of takeout meals during this time. We also report a difference across sex on some of these behavioral changes, with data generally suggesting greater energy intake and less energy expenditure in women than men. Overall, these acute changes in behaviors underscore the significance of a sudden increase in unstructured time at home on potential weight gain.

Concerning eating behavior, as hypothesized, participants increased the consumption of SSBs and pre-packaged processed and ultra-processed foods. Such foods tend to be high in sugar, saturated fat, and sodium content compared to less processed foods and may compose 60-65% of calories of all packaged foods purchased in the U.S.[21]. In addition to having less favorable nutrient content, ultra-processed foods are highly palatable and therefore may produce change in neurocircuitry, causing addictive-like eating behaviors and overconsumption[22, 23]. Not surprisingly, recent research provides fairly consistent support for the association of ultra-processed food intake and SSB intake with excessive weight gain[24, 25]. These processed and ultra-processed foods also have the potential to promote weight gain by altering eating patterns and promoting shifts towards snacking[26]. Our data support this by showing that snacking of calorie-dense savory foods (e.g., chips, popcorn, pretzels, crackers), chocolate, dessert (e.g., cake, cookies, ice-creams, etc.), and yogurt/cheese increased. We speculate that the high energy-density of these snacks along with a possibility that these snacks are typically consumed while engaged in another activity (e.g. while watching TV) may have led to overconsumption. To sum, in line with a recent consumer survey, our participants bought more packaged foods and ate more snacks than usual[27].

The shift towards unhealthy eating was also reflected in the food purchasing behavior where we show that availability and shelf-life of the food, rather than the health, influenced the purchase decisions. While we did not collect the information directly, we speculate that households stocked up on shelf-stable and highly desirable ultra-processed calorie dense foods. Though stocking up on shelf-stable food items is a preparedness necessity, helps minimize trips outside of the home, and is perceived safer than unpackaged foods, we anticipate these purchasing patterns may have contributed to increased energy-dense food in our population group.

In contrast to our hypothesis, our participants reported increasing their intake of fruits and vegetables. Although we did not assess cooking behaviors directly, these findings suggest that spending more time at home may have provided participants the opportunity to cook food at home and utilize more fruits and vegetables in their meals[28]. This speculation is supported by a recent google trends analysis which shows that search for cooking terms increased during the home confinement period, as compared to before pandemic[4]. Notably, participants also reported decreases in the consumption of takeout foods, further suggesting they were likely cooking more. Since takeout foods are often high in calories, total fat, and sodium[29], a decline in the purchase of takeout meals may also have influenced this pattern. Overall, we found that while people were eating more unhealthy foods possibly due to stocking up on shelf-stable foods that are typically highly processed, they also had a high intake of healthy foods possibly due to cooking more meals at home and consuming fewer takeout foods.

Although people reported prioritizing nutrition in their food purchase behaviors, and they did report consuming more fruits and vegetables, they also reported greater intake of energy-dense ultra-processed foods. Their rank-ordering of nutrition as highly important may have reflected a desire for self-presentation or lack of familiarity with the question-type.

Sedentary activities such as sitting, watching television, and other leisurely screen activities all increased significantly with shelter-in-place recommendations. A recent marketing survey reported a similar trend with the use of e-screens in U.S. adults[3]. Evidence regarding the association between sedentary behavior and obesity is weak and inconclusive[30], but it does lead to a decline in energy expenditure. In conjunction with the increased screen time, our participants reported spending less time on vigorous, moderate, and low intensity (walking) exercises. This supports the data from a recent working paper [31], as well as the data released by Fitbit (Fitbit, Inc., 2020) in April 2020, which shows that step count decreased by 16-23% in the younger population. Adults living in dense urban locations are at an even greater disadvantage due to lack of access to outdoor spaces where they can engage in physical activity while maintaining safe social distancing. These variables may further exacerbate the disparity between who can and cannot remain physically active outdoors. Overall, there seemed to be a decline in energy expenditure with a change in PA related behaviors.

Prior studies suggest that women are at higher risk for weight gain than men due to differences in body composition, regulation of food intake, and low total and activity-related energy expenditure[32, 33]. Indeed, women in our dataset reported being more likely to indulge in the consumption of ultra-processed foods, SSBs, energy-dense snacks, as well as fruits and vegetables, than men. Additionally, women were more likely than men to report increases (vs. decreases) in sedentary activities and indulging in screen time. Interestingly, women also reported to have poor sleep quality. Poor sleep quality[34] and sleep deprivation[35] have been associated with abnormal body weight and body fat increases in women in previous literature. Though women increased their sleep time, the possible exposure to high frequency blue screen may have caused disruptions in sleep quality[6], which ultimately may have an effect on body weight. Women also decreased their PA, but not as significantly as men. Overall, as expected, women showed a greater shift towards obesogenic behaviors in our population group. These behaviors also put women at higher risk of weight gain during this pandemic than men.

This study should be considered in light of some limitations. Our sample, though relatively large, was a convenience sample. Moreover, degree of shelter-in-place guidelines and the number of COVID-19 cases in participants area of residence likely differed, creating differences in flexibility with stepping outside the house. Additionally, due to the nature of data collected, our results may be subject to self-reporting bias and/or recall bias. The time frame of data collection may have influenced our results as well. As such, at the time of data collection in early May (April 24^th^ - May 4^th^), although most states had implemented shelter-in-place guidelines, a few states were considering lifting the restriction after May 1^st^. Additionally, this study was cross-sectional, so although associations can be assessed as a result of the pandemic, we cannot establish cause and effect. Finally, we did not use a validated tool for eating behavior measures, or a validated tool for assessing behavior changes, so care should be taken to integrate these findings with the broader literature.

In conclusion, the current findings strongly support the need to implement and support measures that promote strategies to maintain body weight by limiting energy-dense but nutrient poor foods, maintaining healthy eating patterns, performing short bouts of PA, scheduling structured exercises that can be done at home, using tracking applications, and using well-established strategies such as daily self-weighing. Using virtual strategies such as internet-based exercise routines, support groups, virtual activity challenges may be useful to maintain energy expenditure in this the time of extended home confinement. Researchers should also consider collecting body weight data at multiple time points to longitudinally assess the dynamic relationship between behaviors and body weight change. Finally, researchers and public health officials should strategize for ways to reverse any COVID-19-related weight gain or loss of physical fitness that may occur for long-term health.

## Data Availability

Data described in the manuscript (limited to summary data per participant consent), code book, and analytic code will be made available upon reasonable request pending application and approval.

## Acknowledgements

S.B., J.A.C., and M.R.V. conceived and designed the experiment, and acquired the data; M.R.V. analyzed the data and S.B., J.A.C., and M.R.V. interpreted the results; S.B. S.B., J.A.C., and M.R.V. wrote the paper.

